# Widespread transmission in diverse ecotypes challenges visceral leishmaniasis control in East Africa

**DOI:** 10.1101/2025.10.01.25337091

**Authors:** Eva Iniguez, Mercy Tuluso, Steve Kiplagat, Araya Gebresilassie, Esayas Aklilu, Olivia Battistoni, Johnstone Ingonga, John Mark Makwatta, Mohamed Alamin, Osman Dakien, Alphine Chebet, Esther Kaunda, Patrick Huffcutt, Serena Doh, Pedro Cecilio, Galgallo Bonaya, Claudio Meneses, Tiago D. Serafim, Myrthe Pareyn, Mohamed Osman, Eltahir A. G. Khalil, Omran F. Osman, Brima M. Younis, Sithar Dorjee, Guofa Zhou, Jesus G. Valenzuela, Dan K. Masiga, Ahmed M. Musa, Asrat Hailu, Abhay Satoskar, Shaden Kamhawi, Damaris Matoke-Muhia

**Author notes:** Equal contribution (Eva Iniguez, Mercy Tuluso). Corresponding authors: Eva Iniguez; Vector Molecular Biology Section, Laboratory of Malaria and Vector Research, National Institute of Allergy and Infectious Diseases, National Institutes of Health; Rockville, MD 20852, USA. Shaden Kamhawi; Vector Molecular Biology Section, Laboratory of Malaria and Vector Research, National Institute of Allergy and Infectious Diseases, National Institutes of Health; Rockville, MD 20852, USA. Damaris Matoke-Muhia; Centre for Biotechnology Research and Development, Kenya Medical Research Institute, Nairobi 00200, Kenya.

## Abstract

East Africa is emerging as the global hot spot of visceral leishmaniasis (VL), yet efforts to eliminate it are hindered by substantial knowledge gaps in its ecoepidemiology. Here, we report on the high prevalence of *Leishmania* infection in *Phlebotomus orientalis* in Marsabit county, Kenya (3.9%), and Gedaref state, Sudan (3.6%), where this species comprised 99.8% (n = 1185) and 100% (n = 1350) of captured *Phlebotomus* females, respectively. In Aba Roba, Ethiopia, *Ph. martini* accounted for 99% of 184 collected *Phlebotomus* females and had a lower infection rate of 1.5%. *Ph. orientalis* and *Ph. martini* exhibited different habitat and feeding preferences. While *Ph. orientalis* was abundant in diverse peridomestic and sylvatic microhabitats, *Ph. martini* was predominantly collected from termite hills. Moreover, *Ph. orientalis* primarily fed on humans and less on domestic and sylvatic animals. In contrast, *Ph. martini* exhibited zoophagic behavior, mostly feeding on cows and Ovis. Widespread transmission of *Leishmania* in our study sites is supported by high rK39 seroprevalence in both Kenya (17.9%) and Sudan (6.22%). An observed greater prevalence of antibodies to rK39 in individuals living near than away from VL cases in both Kenya (19.8% versus 7.41%, *P =* 0.0015) and Sudan (8.4% versus 2.1%, *P* = 0.0105) demonstrated that proximity to a VL case carries an increased risk of infection. Our findings highlight the need for a risk-based targeted site-specific elimination strategy that accounts for the intensity, diversity, and complexity of VL transmission in today’s East Africa.

## Introduction

East Africa is currently the epicenter of global transmission of visceral leishmaniasis (VL), a fatal neglected disease ranked second after malaria in mortality rates^(1, 2)^. VL in East Africa is epidemiologically and clinically diverse, caused by genetically distinct *Leishmania donovani* strains^(3–5)^, and transmitted by several sand fly vector species including *Phlebotomus orientalis, Ph. martini,* and *Ph. celiae* that have divergent ecologies and bionomics^(6–9)^. VL cases have recently surged in East Africa, emerging in new foci that disproportionally affected Kenya, Sudan, and Ethiopia where they account for more than 65% of cases worldwide, and where ∼50% are children under 15 years old^(1, 10)^.

VL in East Africa had initially been targeted for control rather than elimination due to significant knowledge gaps concerning disease transmission^(11)^. In June 2024, the World Health Organization launched a framework to eliminate VL as a public health concern from East Africa by 2030^(12)^. Elimination is proposed through early diagnosis and treatment, integrated vector control management, and disease surveillance^(12)^. At the country level, the target for elimination was defined as <1% case-fatality rate due to primary VL, 90% decline in new VL cases, absence of death due to VL in children by 2030, and 100% detection and treatment referral of VL-HIV and PKDL patients^(12)^. However, East Africa faces major challenges to VL elimination. Treatment differs between countries, and it is often toxic and lengthy, with low compliance and poor access to care facilities^(6, 12–14)^. In addition, significant knowledge gaps in ecoepidemiology, vector biology, and nature of reservoirs need to be addressed for a successful implementation of an elimination campaign in East Africa,^(6, 13)^ where there are no current effective integrated vector control programs^(12)^. Though *Ph.* orientalis, *Ph. martini*, and *Ph. celiae* sand flies have been found in both peridomestic and sylvatic niches in strong association with *Acacia* and *Balanites* woodlands, vertisol (black cotton soil), and termite hills^(15–18)^, linking these microhabitats to VL transmission in the region^(17, 19–22)^, confirmation of their epidemiological relevance and their potential as resting and breeding sites for these vectors^(18, 23)^ needs to be demonstrated. Additionally, we need to determine the prevalence of asymptomatic individuals and their role as infectious reservoirs for sand flies^(24–26)^. Further hindering control of VL in East Africa is the presence of semi-nomadic populations, such as pastoralists and farmers living in temporary shelters^(13)^, and populations displaced by conflict^(27, 28)^, that are at a higher risk of contracting VL^(27–29)^, complicating the epidemiological landscape.

In this study, we provide a comprehensive account of sand fly species composition, habitat preference, and report the prevalence of *Leishmania*-infected sand flies from diverse microhabitats in endemic study sites in Kenya, Sudan, and Ethiopia. We also compare the prevalence of asymptomatic cases near and away from recently treated VL cases establishing widespread transmission through the high rate of rK39 seroprevalence in Marsabit county, Kenya, and Gedaref state, Sudan, where *Ph. orientalis* is the primary vector. Altogether, this study provides current information on VL transmission dynamics in East Africa.

## Methods

### Ethics approval

The study was approved by the Scientific Research Unit, KEMRI, and licensed by The National Commission for Science, Technology and Innovation (clinical protocol # KEMRI/SERU/CBRD/249/4634) and by the State Ministry of Health and Social Development at Gedaref state, Sudan ethical committee (clinical protocol # UG/EC/3/2023) in Sudan. Both clinical protocols were translated into local languages. Communal consent was obtained by meeting with community leaders. Written consent was obtained from adults or guardians of minors before blood collection. Verbal consent was acquired from homeowners before entomological collections.

### Study sites

We selected study sites with stable VL transmission in Kenya, Sudan, and Ethiopia. The study sites were chosen to encompass features common to VL ecotypes in East Africa.

#### Kenya

Marsabit county has a population of 459,785 people^(30)^, and has experienced sporadic outbreaks since 2014^(31, 32)^. It has an arid and semi-arid tropical savanna climate with an average temperature ranging between 15°C and 26°C. Average precipitation is about 128.24 mm per annum. The long rainy season spans April to May while the short rains occur from November to December; January to March are dry and hot, and June to October are dry cold months ^(33)^. The communal free-range land is dominated by diverse *Acacia* trees and related species, a wide variety of shrubs, and seasonal grass. Inhabitants of Marsabit county are mainly nomadic pastoralists belonging to several tribes, along with traders and farmers, and smaller communities. Camel, goat, sheep, donkeys, and cattle are major domestic animals kept as a source of income. The community moves with their domestic animals from semi-permanent (manyatta) houses to temporary homesteads (foras) for a period of up to six months in search of pasture and water. We investigated Laisamis, Logo Logo, and Karare, sub-counties of Marsabit county.

#### Sudan

Gedaref state has an urban structure with well-established villages. The region is a flat plain, with small, scattered hills and flowing watercourses, and the climate is tropical. The rainy season spans June to October with an annual rainfall of 400-1400 mm. The natural vegetation is a savanna woodland with *Acacia* and *Balanites* trees, and the principal soil type is black cotton vertisol. Agriculture is the main livelihood, with sorghum, sesame, and millet as major crops. Goats, sheep, donkeys, and cattle are major domestic animals kept as a source of income. Most people are subsistence farmers or engage in small animal husbandry. High demand for manual labor attracts seasonal workers from within Sudan or bordering Ethiopia during the rainy season. Dinder National Park falls between the Sahel of Gedaref state and Ethiopian Highlands and is home to several species of large and small mammals, such as lions, gazelle, monkeys, bats and reptiles. A high VL prevalence of 27.6% was recently reported among Dinder National Park wildlife soldiers^(34)^. We investigated the villages of Kunaynah, Sabouni, bordering north Ethiopia, and Umslala-Houng, closest to Dinder National Park, in Gedaref state.

#### Ethiopia

The Konso zone is a semi-arid rugged terrain with many hills, lowlands, and riverbeds. The area is generally semi-arid, with an annual rainfall of about 551 mm per year. The long rains last from February to May, followed by short rains in September and October. The mean annual temperature is 24°C. The predominant soil type is sandy. Dispersed homesteads and animal enclosures are scattered in lowlands or valleys, while villages are on top of large hills or highlands whose slopes lead to the valleys. Segen river flows through the valley providing water for irrigation and creating flat plains suitable for farming and animal herding. Even though there are no permanent human settlements, many families own farms in the valleys and spend a considerable number of nights in temporary shelters (foras) herding cattle, goats, and sheep. The vegetation comprises a mixture of shrub and forests with diverse trees, including species of *Acacia* and *Balanites*. Castellated active or eroded shrinking termite hills^(8)^ are abundant in the area. Aba Roba communities mostly depend on subsistence farming on the cultivated terraced hillsides and flat plains of the Segen River. Sorghum, maize, sesame and sunflower are major crops. Inhabitants also raise goats, sheep, and cattle, kept in enclosures at night. We investigated the villages of Galga and Goinada, 1180-1250 m above sea level, and Maira and Lahalaha, 830-885 m above sea level, in the Konso zone.

### Sand fly collection

In each country, sand fly sampling ecotypes included peridomestic (associated with humans) and sylvatic (away from humans) sampling microhabitats (Supplementary Figure 1). Ecotypes included indoors (inside manyatta/hut) and outdoors (outside manyatta/hut), temporary shelters or foras, animal enclosures, vegetation in association or not with animal burrows and vertisols, and termite hills. The study sites and the total number of CDC light traps used per collection varied in each country, but the same number of traps were placed per ecotype (Supplementary Table 1).

### Identification of *Phlebotomus* sand fly species

The head and the last three segments of each *Phlebotomus* sand fly were wet mounted for morphological identification. Spermatheca and the pharynx teeth were examined to distinguish the female species, while external genitalia were examined for identification of males using a combination of keys^(35, 36)^.

### Rotation of *Phlebotomus* male genitalia

During the first 16-24 h of the adult life of a male sand fly, the external genitalia undergo a rotation to orient the claspers for mating. Thus, unrotated or partially rotated male genitalia can be used to identify newly emerged juveniles, captured during their first night of activity as adults, as a proxy to identify sand fly breeding sites^(15, 18, 20)^. Under the microscope, we recorded the number of males with rotated, unrotated or partially rotated genitalia.

### Mitigation of sample contamination from the field to the bench

In the field, all female sand flies were dissected using dissecting pins that were bleached (10%) and rinsed twice with water before each sand fly dissection. This step was critical to avoid contamination caused by carry-over DNA from sample to sample. In the laboratory, three separate rooms were used to process and run the field samples by qPCR. Room one was used only for preparation of buffer and primer aliquots and loading of the qPCR master mix into the plate. Room two was used for extraction of nucleic acids of field specimens. Room three was used to load the DNA samples into the qPCR plate. Each plate included a standard curve, and DNA from an uninfected midgut and non-template negative controls. To avoid contamination all the instruments, benches and pipettes were continuously bleached (10%) and rinsed with water.

### Extraction of DNA from field-collected *Phlebotomus* female sand flies

Unfed and gravid midgut samples were individually collected in Ethiopia. Individual or pools of 5 midguts were collected in Kenya. For Sudan, pools of 10 sand flies (whole bodies or midguts) were collected. Samples were stored in 50 ml of DNA/RNA Shield buffer (Zymo), and stored at room temperature, or 4°C if available. DNA was extracted from individual specimens of unfed and gravid sand flies using the Quick-DNA/RNA Miniprep Plus kit (Zymo) tissue protocol, with a digestion step of 1 h at 56°C with proteinase K and tissue lysis. DNA was extracted from pools of unfed and gravid sand flies using the Tissue DNA kit (Omega) according to the manufacturer’s instructions. For whole body pooled samples, we first macerated each using a plastic pestle, followed by chemical digestion with proteinase K and tissue lysis buffer at 56°C for 2-6 h or until fully digested.

Blood-fed sand flies were preserved individually as midguts/whole bodies on Whatman 903 Protein Saver cards (Cytica), or in up to 50 ml of DNA/RNA Shield buffer (Zymo). Briefly, field-collected blood fed midguts were individually punched from the Whatman cards using a 3 mm punch biopsy, and Proteinase K (15 ml), Lysis buffer (30 ml), and DNA/RNA shield (30 ml) were added to each punched sample and digested at 56°C for 2 to 4 h, with brief vortexing preformed every hour. Blood fed samples stored in DNA/RNA Shield buffer as midguts/whole bodies were first macerated using ceramic bashing beads (Roche) using the MagNAlyzer (Roche), followed by digestion for 24 h. DNA was extracted using the Quick-DNA/RNA Miniprep Plus kits (Zymo). All samples were eluted to a final volume of 100 µL and stored at −20/-80°C until further processing.

### Blood meal analysis by multiplex PCR

#### Standardization of a blood meal multiplex custom PCR panel

*Lutzomyia longipalpis* sand flies (Jacobina) were reared at the Laboratory of Malaria and Vector Research, NIAID, NIH insectary. Female sand flies (4-6 day old) were allowed to feed for 2 h on whole blood from various species through an artificial chicken skin membrane^(37)^ and collected 24 h post-feeding as positive controls. Blood meals included domestic and sylvatic hosts such as mouse and chicken (LMVR4 animal protocol); human (leftover samples collected for another study under IRB human protocol 000331-I); cow (Charles River Laboratories); dog, goat, sheep, and donkey (Innovative Research); hyrax and camel (donated by Dan Masiga, *icipe*).

A universal reverse primer previously reported^(38)^ was used together with custom forward primers designed in this study based on the mitochondrial *cytochrome b* region, highly conserved among vertebrates^(39)^, to identify several species of interest (Supplementary Table 2). A human primer set based on the cytochrome c oxidase I mitochondrial region^(40)^, and the rodent (*Muroidea* superfamily) primer set targeting the mitochondrial d-loop region reported in previous studies were also used^(41)^. Briefly, DNA (40 ng, 10 μL) was mixed in a single tube as a multiplex reaction with all forward and reverse primers (Supplementary Table 2) and the DreamTaq^TM^ Hot Start Green PCR Master Mix (Thermo Scientific) to a final volume of 30 μL. PCR conditions were 94°C initial denaturation for 5 min, 35 cycles of 30 s at 94°C, 30 s at 55°C, and 60 s at 72°C, and a final extension of 7 min at 72°C. PCR products were separated on a 2% agarose gel electrophoresis ran at 8 v/cm for 60 minutes. Two microliters of a 50 to 1000 base-pair ladder (Thermo Scientific) were used. Bands were visualized on a c600 Azure Biosystems imager.

#### Bloodmeal analysis of field-collected samples

PCR of DNA from field samples was performed using the primers listed in Supplementary Table 2 and following the same procedure and conditions described above. The PCR products were run on a 2% agarose gel. For field samples that did not amplify against any of our custom-made targets, the DNA was amplified using a PCR against generic vertebrate primers (0.2 μM) for the *cytochrome b* gene region^(42)^. The products were separated on a 2% agarose gel and the leftover product was purified (Thermo Scientific) using the GeneJet PCR purification kit (Thermo Scientific), and reamplified under the same conditions. After confirming amplification by agarose gel, the remaining reamplified product was purified and sequenced (Eurofins Genomics). Sequences were aligned with reference genomes from the NCBI database to determine blood meal identity.

### *Leishmania* detection and quantification by qPCR

A standard curve was created for each different sample condition by spiking and uninfected sand fly midgut, a whole body, or a pool of whole bodies, with 10^5^ *L. donovani* (MHOM/SD/62/1S) parasites followed by DNA extraction, followed by a 10-fold serial dilution into corresponding uninfected samples.

For unfed and gravid samples (processed individually or as a pool of 5-10 sand flies), we used qPCR to amplify the kinetoplast minicircle from *Leishmania* parasites using JW11 and JW12 primers^(37)^ in a 25 µL reaction volume containing SYBR green (Perfecta SYBR Green Fast Mix or FIREPol Master Mix Ready to Load). PCR conditions were 95°C initial denaturation for 3 min with 49 cycles of 10 s at 95°C and 30 s at 55°C. A probe-based qPCR was used for individually blood fed sand flies as described^(37)^. All field samples were run in duplicate (unfed/gravid) or triplicate (blood fed) using 40 ng of DNA template. A standard curve of serially diluted parasite-spiked midguts, negative uninfected laboratory-reared sand flies, and positive *Leishmania donovani*-infected laboratory-reared sand flies were included as controls in each plate. The melting curve of field-samples was compared to the melting curve of positive and negative controls. Samples considered positive for *Leishmania* kDNA by qPCR were re-run to confirm positive amplification.

### Human dried blood spot collection

All reported VL cases to the Ministry of Health were diagnosed and treated at reference hospitals according to national guidelines^(12, 43)^. Males and females were included in our study. A total of 752 participants were enrolled from Laisamis and Karare villages, Marsabit county, Kenya, and 435 participants were enrolled from Kunaynah and Sabouni villages, Gedaref state, Sudan. Blood was collected by finger prick using a lancet, spotted on a Whatman 903 protein saver card (Cytiva), and preserved at room temperature. For Kenya, non-neighboring controls lived in a neighborhood located within 5 km from areas of index VL cases within Laisamis but with no reports of VL cases. In Sudan, controls households were located in Sabouni, an adjacent village at a distance of 80 km from Kunaynah, with no reports of VL cases, a similar ecology, and where sand flies are present.

### *Leishmania* rK39 ELISA from dried blood spots

ELISA plates (Thermo Fisher Scientific) were coated overnight at 4°C with 25 ng/well of rK39 antigen (donated by Steve Reed from the Infectious Disease Research Institute, now the Access to Advanced Health Institute). In parallel, dried blood spots (DBSs) were eluted with 300 μl of PBS 0.05% Tween 20 (Sigma) and incubated for 4 h shaking at room temperature or overnight at 4°C. Plates were blocked with 200 μl diluted in Tis-Bris Saline 0.05% Tween 20 (TBST) with 20% donor equine serum (DES; Cytiva), and incubated for 1 h at room temperature. After blocking, 25 μl of the eluted DBS sample was diluted in TBST/5% DES (1:4 dilution) to a total volume of 100 μl/well in duplicates. Plates were incubated for 1 h at room temperature, followed by 6 washes with TBST. One hundred microliters of secondary antibody, [Goat anti-human IgG (H+L) Alkaline phosphatase conjugated; Sigma], diluted in TBST/5% DES was added to each well. After 1 h of incubation, the plates were washed 12 times with TBST. Lastly, 100 μl of substrate-p-nitrophenyl phosphate liquid (Sigma) was added to each well and data was collected 1 h after addition of the substrate at a 405 nm wavelength. Non-endemic negative controls obtained from individuals living in Nairobi, Kenya were included in each plate for normalization.

### Statistical analysis

A chi-square goodness-of-fit test was performed for the entomology data, followed by post-hoc pairwise comparisons using standardized residual (Z-score) analysis with Bonferroni correction to identify differences among ecotypes within each country. A Bonferroni-adjusted critical value of |Z| 2.69 (corresponding to *P* value ≤ 0.007) was considered significant. ELISA data was normalized using the plate cutoff calculated by the mean optical density (O.D.) of non-endemic negative controls + 5 SD, followed by log transformation. After normalization, samples were statistically compared using the Mann-Whitney test. Odds ratio was used to assess the risk for having rK39 antibodies (categorized as positive or negative) between endemic index case household members and neighborhood households versus non-neighboring households (Kenya), or non-neighborhood controls (Sudan), and its significance was tested using Fisher’s exact test. A univariable binary logistic regression was performed to assess the effect of distance in meters from index case households to neighboring versus non-neighboring households for Kenya, or distance in meters from a VL case in Kunaynah to households a non-endemic adjacent village for Sudan, on rK39 seropositivity (risk by log-odds of positive outcome after normalization). The ‘margins’ command of Stata was used to compute adjusted log-odds of seropositivity at selected distances from index case households. A *P* value of ≤ 0.05 was considered significant.

## Results

### Study sites in Kenya, Sudan, and Ethiopia

Figure 1 depicts our selected endemic study sites in Kenya, Ethiopia and Sudan (Figure 1A) and their stable VL transmission (Figure 1B). Marsabit county in Kenya experienced sporadic outbreaks^(31, 32)^, reporting an incidence of 1 case per 10,0000 inhabitants over the past seven years excluding a notable increase in incidence during 2019. The Konso zone in Ethiopia, reported 2.66 cases per 10,000 inhabitants per year since 2018. In Sudan, Gedaref state is a highly endemic VL region that borders Ethiopia to the east and reports a steady 10 cases per 10,000 inhabitants per year since 2017 (Figure 1B).

**Figure 1.**
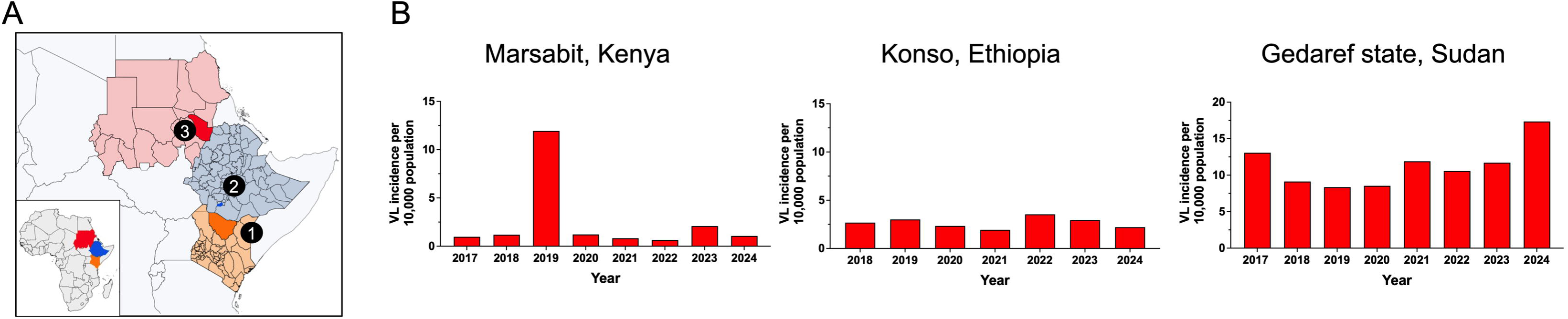
Study sites and incidence of visceral leishmaniasis. (A,B) Map of East Africa showing the location of study sites. 1, Marsabit; 2, Konso; 3, Gedaref state. (B) Visceral leishmaniasis (VL) cases per 10,000 population over the past seven years in Marsabit county, Kenya; Konso, Ethiopia; and Gedaref state, Sudan. Incidence data are based on Ministry of Health reports from each country. Maps at QGIS software QGIS shapefile, https://open.africa/dataset/?tags=Shapefiles.

### Distinct microhabitat and breeding preferences of vector species

Entomological surveys of phlebotomine sand flies were carried out periodically from September 2023 to September 2024 (Supplementary Table 1). Trapping sites were selected from representative peridomestic and sylvatic ecotypes (Supplementary Figure 1). Our collections determined that *Ph. orientalis* was the predominant VL vector species in Marsabit, Kenya, and Gedaref, Sudan, comprising 99.75% (n = 1185) and 100% (n = 1350) of *Phlebotomus* females, respectively (Figure 2A). In contrast, *Ph. martini/Ph. celiae* females (morphologically indistinguishable) were the most prevalent of five *Phlebotomus* species in Konso, Ethiopia, accounting for 99% of the collection (n = 184; Figure 2A). Other female *Phlebotomus* species captured included one *Ph. martini/Ph. celiae* and two *Ph. alexandri* in Kenya; and six *Ph. orientalis*, four *Ph. saevus,* and one *Ph. alexandri* in Ethiopia.

**Figure 2.**
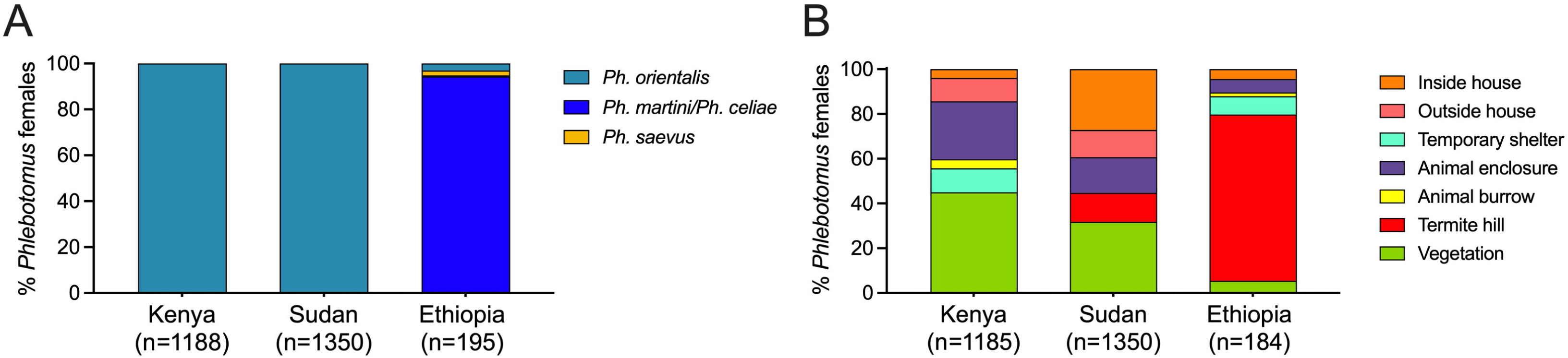
Distribution of different sand fly species across distinct microhabitats in endemic foci in Kenya, Sudan and Ethiopia study sites. (A) Relative abundance of collected *Phlebotomus* females by species. (B) Relative abundance of *Phlebotomus* females by ecotype.

Comparing ecotype productivity of *Ph. orientalis* and *Ph. martini/Ph. celiae* revealed a significant difference in habitat distribution across all study sites in the three countries (c^2^ = 1352.89, d.f. = 12, *P* <0.0001). The majority of *Ph. orientalis* females in Marsabit, Kenya, were collected from outdoor habitats associated with peridomestic and sylvatic vegetation (45.01%, *P* <0.0001), mostly *Acacia* trees, followed by animal enclosures (25.92%, *P* <0.0001; Figure 2B). In Gedaref, Sudan, *Ph. orientalis* was primarily captured in vegetation (31.78%, *P* <0.0001) and inside houses (27.19%, *P* <0.0001) whose floors commonly consist of black cotton soil, a risk factor for VL^(17, 22)^. The remaining specimens were collected from outside houses (12.15%) and in animal enclosures (15.93%; Figure 2B). Worth noting that in Sudan, the majority of *Ph. orientalis* females (39.34%) were collected near human dwellings. In Konso, Ethiopia, *Ph. martini/Ph. celiae* females were mainly captured from termite hills (74.46%, *P* <0.0001) that were prevalent in both peridomestic and sylvatic ecotypes, followed by temporary shelters locally known as “foras” (8.15%; Figure 2B).

We used *Phlebotomus* males to verify species prevalence and to indicate breeding sites. *Ph. orientalis* represented 100% of all collected males in Kenya (n *=* 260) and Sudan (n = 4714) (Supplementary Figure 2A). Like females, *Ph. orientalis* males in Marsabit, Kenya, were collected from vegetation (46.92%, *P* <0.0001), outside houses near *Acacia* and *Balanites* trees (18.85%), and in animal enclosures (19.23%) (Supplementary Figure 2B). Preliminary evidence of breeding sites, identified by finding unrotated or partially rotated male genitalia (n *=* 12), implicated outdoor ecotypes (Supplementary Figure 2C). In Gedaref, Sudan, *Ph. orientalis* males were mainly collected from vegetation (33.64%, *P* <0.0001) and indoors (22.70%, *P* <0.0001) (Supplementary Figure 2B), similar to females. Of the captured males with unrotated or partially rotated genitalia (n *=* 33), 51.52% (*P* <0.0001) were collected from vegetation and 21.21% from animal enclosures (Supplementary Figure 2C), potentially indicating that *Ph. orientalis* may not be breeding indoors in either Kenya or Sudan.

In Konso, Ethiopia, *Ph. martini* was the main male species collected (Supplementary Figure 2A), mostly from termite hills (72.35%, *P* <0.0001) (Supplementary Figure 2B), again similar to females. Other male species included *Ph. celiae* (5.80%) and *Ph. orientalis* (1.02%) (Supplementary Figure 2A). Some 72.92% (n *=* 35, *P* <0.0001) of *Ph. martini* males with unrotated or partially rotated genitalia were collected from termite hills (Supplementary Figure 2C), implicating this microhabitat as a significant breeding site for *Ph. martini*. Of note, we collected one *Ph. celiae* male with unrotated/partially rotated genitalia from a domestic animal enclosure. The relative abundance of *Ph. martini* (94.14%) over *Ph. celiae* males (5.80 %) indicates predominance of the former species in our female collections. As such, we will use *Ph. martini* instead of *Ph. martini/Ph. celiae* for females from here on.

### Vector species exhibit different feeding preferences

We performed host blood meal analysis on field-collected individual blood fed *Phlebotomus* females using a custom-made multiplex PCR tailored to amplify blood from humans and seven animals present in our study sites (Supplementary Figure 3, Supplementary Table 2). The multiplex PCR panel identified 63.8% (n = 270/423) of blood meals. After *cytochrome b* (*cytb*) gene sequencing of unknowns this increased to 96.9% (n *=* 410/423). Interestingly, the diversity of identified blood meal sources was considerably higher in Marsabit, Kenya, (13 hosts) compared to either Gedaref, Sudan, (5 hosts) or Konso, Ethiopia, (4 hosts) (Figure 3A-C). Of interest, the observed number of sand flies with multiple blood meals were higher than expected for Marsabit, Kenya, (c^2^ = 44.09, d.f. = 15, *P* <0.0001) and Gedaref, Sudan (c^2^ = 17.83, d.f. = 6, *P* <0.0067). Our results also establish that several females contained mixed blood meals from humans and domestic or sylvatic animals such as hyraxes and mongooses (Figure 3A-C).

**Figure 3.**
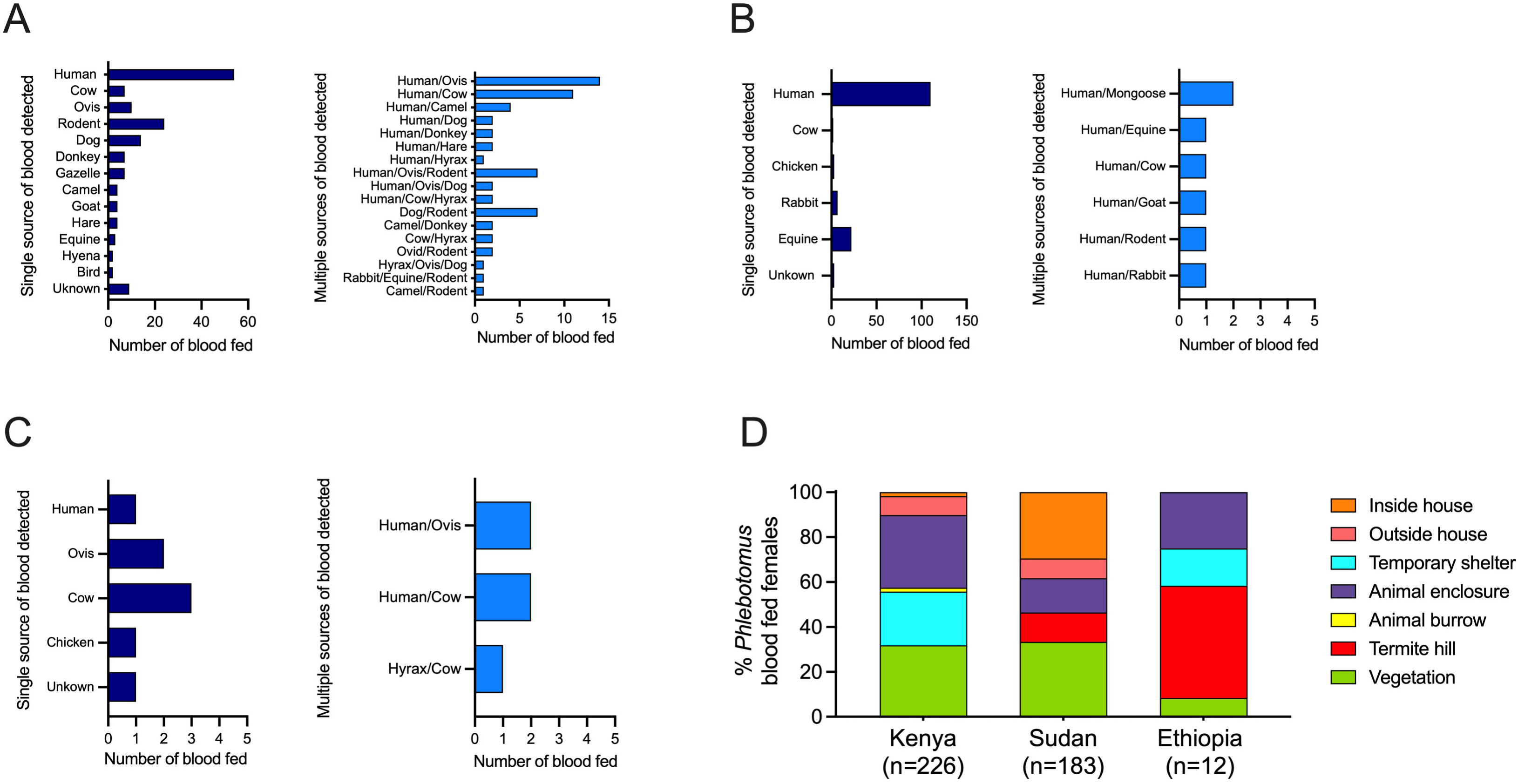
A mosaic of blood meal sources observed in leishmaniasis vectors across endemic foci in Kenya, Sudan and Ethiopia study sites. (A-C) Host blood meal identity of blood fed females in Kenya (A), Sudan (B), and Ethiopia (C) with a single or multiple sources of blood detected. (D) Relative abundance of collected blood fed *Phlebotomus* females by ecotype.

In Marsabit, Kenya, we collected 226 *Ph. orientalis* and two *Ph. alexandri* blood fed females from animal enclosures (32.30%, *P* <0.0001), vegetation (31.86%, *P* <0.0001), and temporary shelters (23.89%, *P* = 0.001), following a similar pattern to unfed/gravid sand flies (Figure 3D). Humans were identified as the dominant host for *Ph. orientalis*, representing 37.76% (*P* <0.0001) of single blood meals, followed by rodents (15.89%, *P* <0.0001), dogs (9.27%) and other sylvatic animals like gazelles (4.64%) (Figure 3A). Mixed blood meals from two or three hosts were identified in 63 *Ph. orientalis* specimens including human/Ovis (22.22%), and human/cow (17.46%) (Figure 3A). In Gedaref, Sudan, 29.51% (*P* <0.0001) of the 183 collected blood fed *Ph. orientalis* specimens were captured inside the house, and 33.33% (*P* <0.0001) in vegetation (Figure 3D). Similar to Marsabit, Kenya, humans (74.83%, *P* <0.0001) were the predominant host, while 14.97% fed on donkey (Figure 3B). Only seven specimens contained mixed blood meals that fed on human plus other domestic and sylvatic animals such as mongooses and rabbits (Figure 3B). In Ethiopia, only 12 *Ph. martini* blood fed specimens were collected from termite hills (50%, *P* <0.001) and animal enclosures (25%) (Figure 3D). Contrary to the anthropophilic feeding behavior observed for *Ph. orientalis*, *Ph. martini* seemed to preferentially feed on cows (37.5%), and Ovis (25.0%), and five sand flies fed on multiple hosts such as human/cow and hyrax/cow (Figure 3C). Of the few collected *Ph. alexandri* blood fed females, the two from Kenya fed on humans, while in Ethiopia, the one specimen fed on an Ovis.

### *Leishmania-*infected vector species were recovered from diverse microhabitats

Using qPCR (Supplemental Figure 4) on individual (n_i_) or pooled (n_p_) sand flies, we detected *Leishmania*-infected specimens in both peridomestic and sylvatic niches across all study sites (Figure 4A), and from distinct ecotypes (Figure 4B-C). The minimal infection rates of *Ph. orientalis* with *Leishmania* were similar in Marsabit, Kenya, (26/648, 3.9%) and Gedaref, Sudan, (10/281, 3.6%). Ethiopia had a lower rate of infection of 1.5% (3/183) for *Ph. martini* females (Figure 4A). Of note, none of the other sand fly species collected throughout this study were infected.

**Figure 4.**
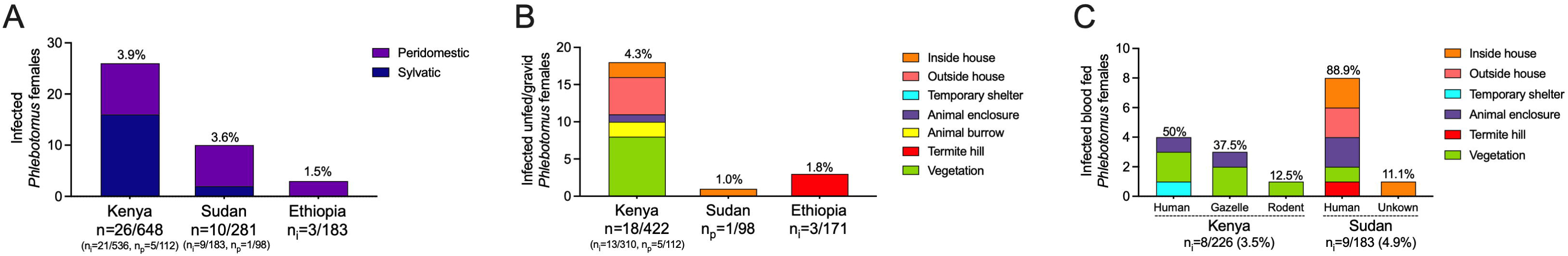
Distribution of *Leishmania*-infected sand flies from distinct and diverse microhabitats reveals hotspots of active transmission near and away human habitation. (A) Number of *Leishmania*-infected sand flies collected from peridomestic or sylvatic habitats. (B) Habitat productivity of infected unfed/gravid female *Ph. orientalis* sand flies in Kenya and Sudan, and *Ph. martini/Ph. celiae* in Ethiopia. (C) Host blood meal identity of infected blood fed *Ph. orientalis and Ph. martini/Ph. celiae* females. Specimens were processed individually or as pools of up to 5-10 sand flies (B) or individually (C).

In Marsabit, Kenya, 4.3% (18/422) unfed/gravid and 3.5% (8/226) blood fed *Ph. orientalis* were *Leishmania* positive (Figure 4B-C). Infected *Ph. orientalis* were associated with vegetation (30.77%), where we captured ten infected unfed/gravid and four infected blood fed females in sylvatic sites, and seven unfed/gravid and three blood fed specimens from peridomestic sites (Supplementary Tables 1 and 3). Interestingly, a cluster of six infected sand flies was collected outside and inside the house of a relapsed VL patient in Laisamis, Kenya (Supplementary Tables 1 and 3), alluding to how hotbeds of transmission form. The patient, negative for the human immunodeficiency virus, received treatment in 2021 and refused treatment after relapsing in January 2024 (Supplementary Table 3), highlighting the need for better surveillance and treatment. Though four of the eight (50%) infected blood fed *Ph. orientalis* fed on humans, confirming the vector’s anthropophilic behavior, finding three and one infected sand flies that fed on gazelles and a rodent, respectively (Figure 4C, Supplementary Tables 3 and 4), stresses the need to investigate whether there is a role for animal reservoirs in VL transmission in East Africa.

For Gedaref, Sudan, one of the 98 (1.0%) screened unfed/gravid specimens collected from inside a house was positive for *Leishmania* (Figure 4B). Importantly, we found nine (9/183, 4.9%) blood fed *Leishmania*-infected *Ph. orientalis* females, seven from peridomestic sites, and two from vegetation and a termite hill in sylvatic sites within Dinder Park (Figure 4C, Supplementary Table 3). All nine infected sand flies fed on humans, further emphasizing the anthropophilic behavior of *Ph. orientalis* in this region (Figure 4C, Supplementary Tables 3 and 4). The focal nature of transmission was again revealed by finding three infected blood fed sand flies, two inside the house of a VL patient, collected in consecutive days, and a third that was captured from inside the house of the patient’s neighbor (Supplementary Tables 1 and 3).

In Ethiopia, three (1.8%, n = 3/171) unfed/gravid *Ph. martini* were positive for *Leishmania,* all captured in termite hills (Figure 4B). Two of the three were captured 9 months apart from the same castellated termite hill located in Galga near the house of a recent VL case (Supplementary Tables 1 and 3), again highlighting the focality of VL. The prevalence of *Ph. martini* females in termite hills and the exclusive collection of *Leishmania*-infected specimens from this microhabitat implicates it in VL transmission in Ethiopia, and potentially elsewhere in East Africa.

### *Leishmania* rK39 seroprevalence reveals an increased risk of infection for individuals living near VL cases

As *Ph. orientalis* is the vector of *L. donovani* in both Marsabit, Kenya, and Gedaref, Sudan, we assessed seroprevalence of *Leishmania* rK39 antibodies in these study foci. We conducted a case-control study of household members of VL cases and five neighboring households for comparison against non-neighboring controls. These comprised households located approximately 1.5 to 10 km from respective index VL cases within the endemic villages (Kenya) or households located in an adjacent non-endemic village 80 km away from Kunaynah (Sudan). DBSs were collected from household members of 48 VL cases diagnosed during December 2023 to May 2025 in Laisamis and Karare villages, Marsabit county, Kenya, and household members of five VL cases diagnosed during August 2023 to August 2024 in Kunaynah village, Gedaref state, Sudan (Supplementary Table 5). In Kenya, we enrolled a total of 752 subjects (male to female ratio of 1:1.54) with a median age of 14 years (range, 2 – 85 years). In Sudan, we enrolled a total of 435 participants (male to female ratio of 1:1.3) with a median age of 15 years (range, 5 months – 85 years). Thirty-two (n = 108 household members) and ten (n = 145 household members) control households were screened in Kenya and Sudan, respectively.

rK39 titers were significantly higher for individuals living with or near cases (neighboring households) than for those of non-neighboring controls in both Kenya and Sudan (Figure 5 A-B). In Kenya, 19.80% (118/596) of screened asymptomatic participants living with or near VL cases exhibited antibodies against rK39, compared to 7.41% (8/108) of those living farther away. Similarly, 8.42% (24/285) of asymptomatic people living with or near VL cases in Sudan were seropositive, versus only 2.07% (3/145) of those living away from cases. The odds ratio calculated by Fisher’s test for those living with or near a VL case was 3.08 (*P* = 0.0015) and 4.35 (*P* = 0.0105) for Kenya and Sudan, respectively. Additionally, logistic regression demonstrated that the likelihood of developing rK39 antibodies decreases significantly with distance from a VL case for both Kenya (Figure 5C; Likelihood ratio = 19.8805, *P* < 0.0001; z = −3.1344, *P* = 0.0017) and Sudan (Figure 5D; Likelihood ratio = 7.2247, *P* = 0.0072; z = −2.2891, *P* = 0.0221). Importantly, the same outcome was observed when the analysis excluded VL index case household members for both Kenya (Supplemental Figure 5 A; Likelihood ratio = 16.5773, *P* < 0.0001; z = −2.95592, *P* = 0.0031) and Sudan (Supplemental Figure 5B; Likelihood ratio, 5.71154, *P* = 0.0169; z = −2.08351, *P* = 0.0372). Of note, at the time of serum collection, only 22/48 (Kenya) and none (Sudan) of the diagnosed and treated VL patients had detectable antibodies by rK39 ELISA (Supplementary Table 5), considered the gold standard VL serodiagnostic test, even though DBSs were collected within 1-4 months of treatment.

**Figure 5.**
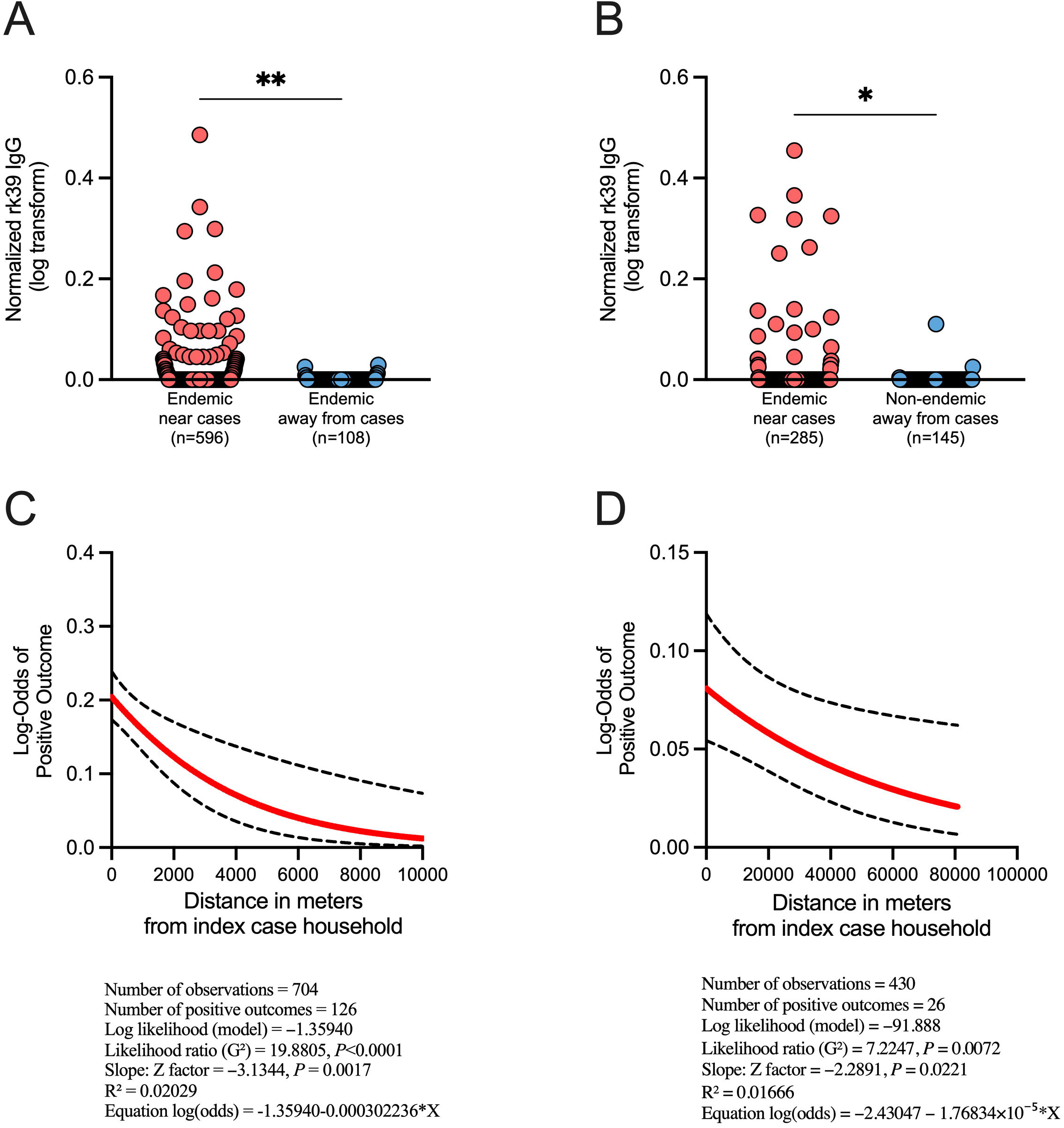
Prevalence of *Leishmania* rK39 antibodies in our study sites indicate high risk of infection in Marsabit county, Kenya, and Gedaref state, Sudan. (A,B) *Leishmania* rK39 antibody prevalence measured from dry blood spots (DBS) collected from individuals living with or near and away from recently reported VL cases in Laisamis and Karare, Marsabit county (A) and Kunaynah, Gedaref state (B). Data was normalized using the plate cutoff calculated by the mean optical density (O.D.) of non-endemic negative controls + 5 SD. Non-endemic negative controls were obtained from individuals living in Nairobi, Kenya. Samples with O.D. ≥ 0.00 after normalization were considered positive. (C,D) Univariable binary logistic regression analysis was performed to assess the effect of distance (meters) from index case households to neighboring and control households, on rK39 positivity in Marsabit, Kenya (C), and Gedaref, Sudan (D). The ‘margins’ command of Stata was used to compute adjusted log-odds of seropositivity at selected distances from the index households. Analysis includes index household members. A *P* value of ≤ 0.05 was considered significant by Mann-Whitney test (A,B), or univariable binary logistic regression (C,D); \**P* < 0.05, \*\**P* < 0.01.

## Discussion

The WHO has recently targeted visceral leishmaniasis in East Africa for elimination as a public health concern by 2030^(12)^. Our findings provide evidence of widespread and intense VL transmission in diverse settings across our study sites to inform control efforts.

Consistent with earlier reports from the region^(7, 9, 44)^, *Ph. orientalis* was the most prevalent *Phlebotomus* species in Sudan and northern Kenya, while *Ph. martini* predominated in southern Ethiopia. We established that *Ph. orientalis* was predominant in *Acacia* forests in association with vertisol and animal burrows, with a high proportion of juvenile males collected outside, in vegetation and animal enclosures, demonstrating that these microhabitats are suitable breeding sites for this vector as previously suggested^(9, 15, 18, 45)^. In comparison, termite hills were the most productive site for *Ph. martini*, as reported previously^(8, 46–48)^, and the collection of over 72% of newly emerged males, provides direct evidence of this ecotype as a preferential breeding site for *Ph. martini*.

We recovered *Leishmania*-infected *Ph. orientalis* from different microhabitats within our study sites in Marsabit, Kenya, and Gedaref, Sudan, in both sylvatic and peridomestic niches, and frequently from the same location months apart. This indicates that *Ph. orientalis* has adapted to diverse ecological microhabitats that provide sugar and blood meal sources, and breeding habitats^(15, 16, 18)^, driving widespread yet focal transmission of VL. Supporting this observation, our data indicates that *Ph. orientalis* likely shifts its behavior to adapt to climatic conditions. Collections from Helat-Belo, Gedaref state, Sudan, conducted during the dry season in March to June 2016-2018 found infected blood fed *Ph. orientalis* mainly outside houses and in sylvatic ecotypes, indicating they are exophilic, and reported that they fed on human, cattle, and donkeys^(17)^. In contrast, our sand fly collections from Kunaynah and Umslala-Houg were performed during the rainy season (June to September), when people tend to sleep inside their houses. Consequently, *Leishmania*-infected *Ph. orientalis* females were collected mainly indoors and had fed soley on humans, suggesting a potential shift in vector behavior, with sand flies likely resting inside the house after feeding on humans. While the majority of juvenile *Ph. orientalis* males were collected from vegetation and animal enclosures, we also collected juvenile males indoors in Sudan, possibly due to the widespread nature of vertisols (black cotton soil) in the village, including inside houses, serving as a breeding site for this vector^(18)^, and creating an ideal environment for focal and active VL transmission indoors. Further confirmation of this shift in vector behavior and transmission pattern is needed to inform control measures that are currently focused on outdoor applications.

In Konso, Ethiopia, *Ph. martini* were predominantly collected from termite hills, both in sylvatic and peridomestic settings, pointing to a restricted habitat preference compared to *Ph. orientalis*. Additionally, all three *Leishmania*-infected sand flies were recovered from termite hills reinforcing findings of a previous study^(8)^. Capturing two *Leishmania*-infected females nine months apart from a peridomestic castellated termite hill reinforces the focality of infection sources and highlights the importance of this microhabitat in sustaining transmission by *Ph. martini*. Notably, this termite hill was located only 20 meters away from a household that reported a VL case. In-depth studies focused on termite hills are needed to fully elucidate the involvement of this microhabitat in VL transmission.

Several studies have reported infection rates in field-collected sand flies from Kenya (0.3%), Sudan (1.6% and 1.4%)^(17, 49)^, and southern (5.8%)^(8)^ and northern Ethiopia (23%)^(21)^. In our study, infection rates were higher than was previously reported for Kenya (3.9%) and Sudan (3.6%), and were lower for southern Ethiopia (1.5%). Additionally, reinforcing previous findings, we recovered infected specimens from both sylvatic and domestic/peri-domestic habitats^(8, 17, 19, 21, 49, 50)^.

Host attractiveness to vector sand flies, seroprevalence of *Leishmania* antibodies, and PCR detection of *L. donovani* DNA have been reported for various domestic and wild animal species in East Africa^(51–55)^, but live parasites have not been isolated from any thus far. Irrespective of whether domestic or wild animals are reservoirs, VL endemicity in scarcely populated areas supports the existence of a zoonotic transmission cycle in East Africa^(17, 22, 56)^. Establishing whether animal reservoirs are involved in sustaining VL transmission in East Africa is a major knowledge gap that needs to be urgently addressed to inform on the best strategy for VL elimination in this region.

Finding *Leishmania*-infected sand flies near VL cases from all our study sites affirms the importance of human reservoirs in maintaining VL transmission, and the need for continued surveillance and rapid treatment of cases. This is further reinforced by rK39 serological surveys where participants living near VL cases exhibited a significantly higher proportion of positive rK39 antibodies and an increased risk associated with proximity to VL cases (log-odds of positivity decreased with increasing distance from the index households) in both Marsabit, Kenya, and Gedaref, Sudan. Importantly, the high prevalence of rK39 antibodies in asymptomatic participants provides further evidence of prevalent and efficient transmission, and stresses the urgency of determining their involvement as potential parasite reservoirs^(57)^.

A limitation of our study is the use rK39 to assess seroprevalence and the small sample size of VL index cases in Sudan. The sensitivity of rK39 in East Africa is known to be suboptimal^(58–60)^. In our study, this is reinforced by the loss of rK39 positivity in recently diagnosed VL cases emphasizing the need for diagnostic tools with better specificity and sensitivity to detect *Leishmania* infections in the region. Although we observed a significant decrease in rK39 seropositivity with increasing distance from index VL case households in both Kenya and Sudan, future studies with a larger sample size are needed to assess the increased risk of living in proximity to VL cases. For studies of vector sand flies, our aim was to identify their preferred habitats through a broad investigation of distinct foci within three East African countries. Moving forward, we need in-depth longitudinal studies focused on identified hotbeds of infection to better understand the requisites supporting transmission in specific microhabitats.

Our exploratory ecoepidemiological study conducted in three East African countries reports widespread VL transmission in the region and emphasizes both its focality and complexity. The information we provide on habitat preference and transmission hotspots of vector species in distinct microhabitats will galvanize further studies across disease foci throughout East Africa, needed for an effective elimination campaign.

## Acknowledgements

We thank the community, administrative officers, and health care workers from all our study sites. We are also grateful to Marsabit County, Kenya, for supporting us with fieldwork implementation and logistics. We thank Professor EL-Hassan Center for Tropical Diseases staff, community leaders and the population in the Kunaynah village, Gedaref state, Sudan. We thank the Karat Zone Health Bureau, Ethiopia, for supporting this work and the Arbaminch University for hosting the project in Southern Ethiopia. We also acknowledge Rogers Asamba (KEMRI), and David Mbuvi (ICIPE) for their valuable support during fieldwork; Dr. Geyeto Garra (Ethiopia) for support given to this project; Mr. Haile Gebremariam (Ethiopia) for technical assistance.

## Author contributions

E.I. performed the experiments, analyzed the data, conceived the study, supervised the work, and wrote the manuscript. M.T., Steve Kiplagat, A.G., E.A., O.B., J.I., J.M.M., M.A., O.D., A.C., E.K., P.H., S.D., P.C., G.B., C.M., T.D.S., M.P., M.O., E.A.G.K., O.F.O., and B.M.Y. and D.M.M. undertook the field work, performed the experiments, analyzed the data, and edited the manuscript. G.Z. and S.D. performed the sand fly statistical analysis. J.G.V., D.K.M., A.M.M., A.H., A.S., E.I. and Shaden Kamhawi wrote the manuscript. E.I., J.G.V., D.K.M., A.M.M., A.H., A.S., Shaden Kamhawi, and D.M.M. conceived the study and supervised the work.

## Data availability

All data produced in the present work are contained in the manuscript

## Declaration of Generative AI and AI-assisted technologies in the writing process

During the preparation of this work, the authors used GPT-4o (OpenAI, NIAID GenAI Toolkit) in order to improve readability and language of the manuscript. After using this tool/service, the authors reviewed and edited the content as needed and take full responsibility for the content of the publication.

