## Supplemental file for "Widespread transmission in diverse ecotypes challenges visceral leishmaniasis control in East Africa"

[†]Equal contribution

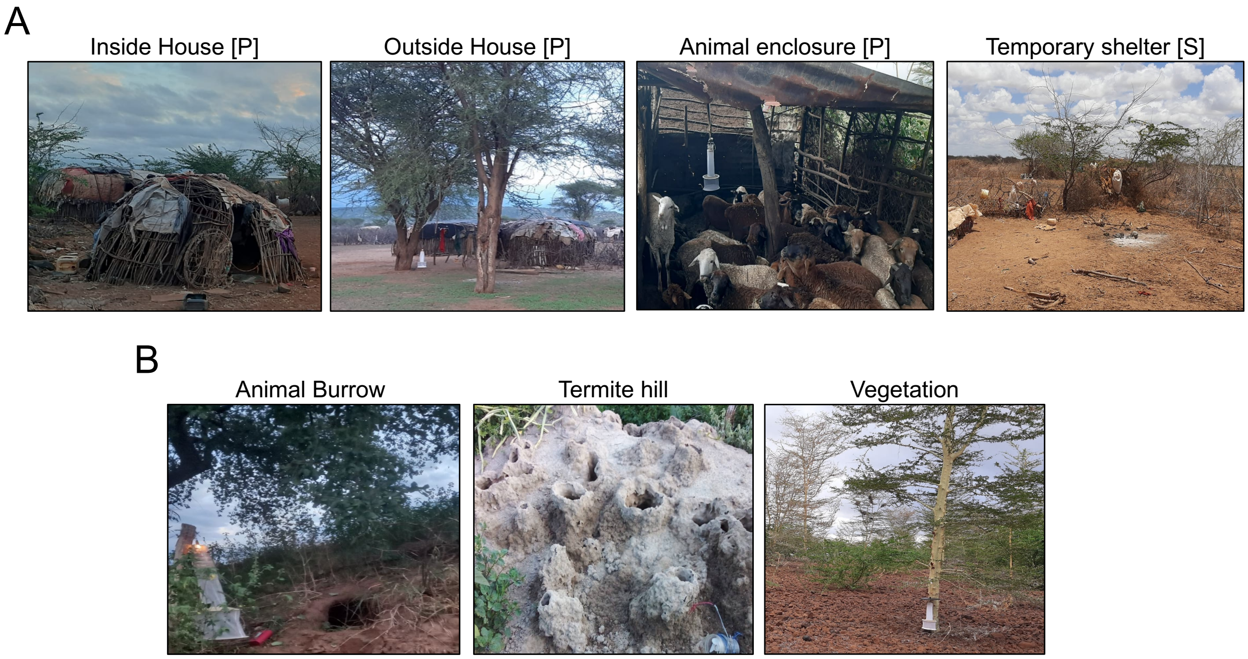


**Supplemental Figure 1. Peridomestic and sylvatic sand fly trapping ecotypes**
(A) Representative images of ecotypes present in peridomestic (P) or sylvatic (S) sites. (B) Representative images of ecotypes present in both peridomestic and sylvatic sites.

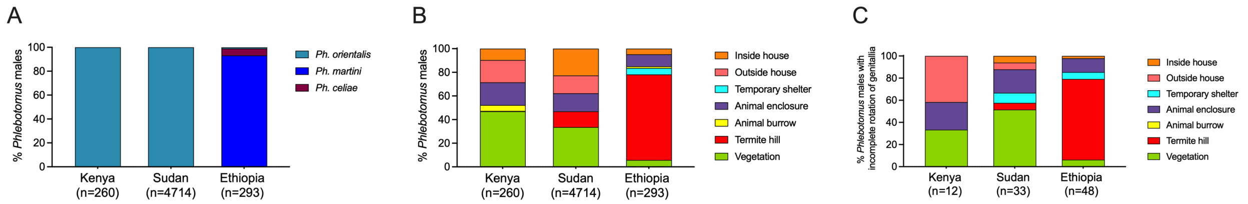


**Supplementary Figure 2. Males reveal the breeding microhabitats of VL vectors in East Africa**

(A) Relative abundance of collected *Phlebotomus* males by species. (B) Relative abundance of *Phlebotomus* males by ecotype. (C) Breeding habitat productivity based on finding unrotated/partially rotated male genitalia.

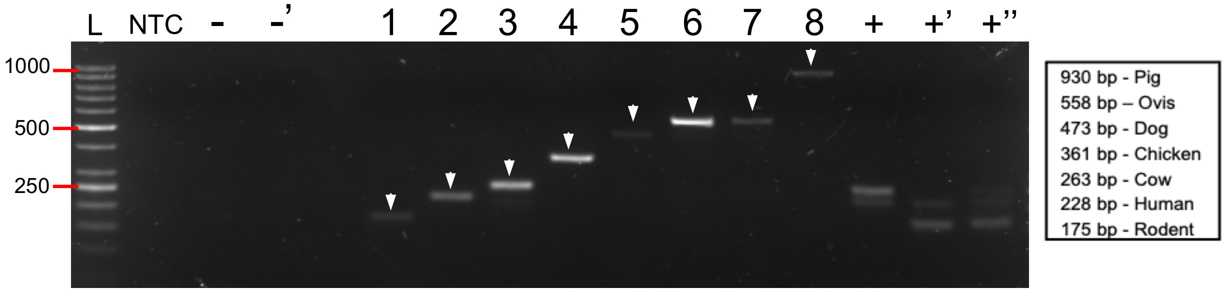


**Supplementary Figure 3. Representative PCR panel for multiplex blood meal analysis.** Multiplex PCR panel for blood meal analysis. Gel shows bands of distinct sizes to distinguish human and seven common vertebrate hosts in East Africa. 1, rodent; 2, human; 3, cow; 4,chicken; 5, dog; 6, goat (Ovis); 7, sheep (Ovis); 8, pig. Negative controls, DNA from unfed sand fly (-), or from an experimental sand fly fed on rabbit blood absent in this panel (-’); Positive controls, DNA from an experimental sand fly fed on a mixed cow/human (+), rodent/human (+’), rodent/cow/human (+”) blood meals. L, ladder. NTC, non-template control. Arrowhead, single blood meal. Rectangles, mixed blood meals.

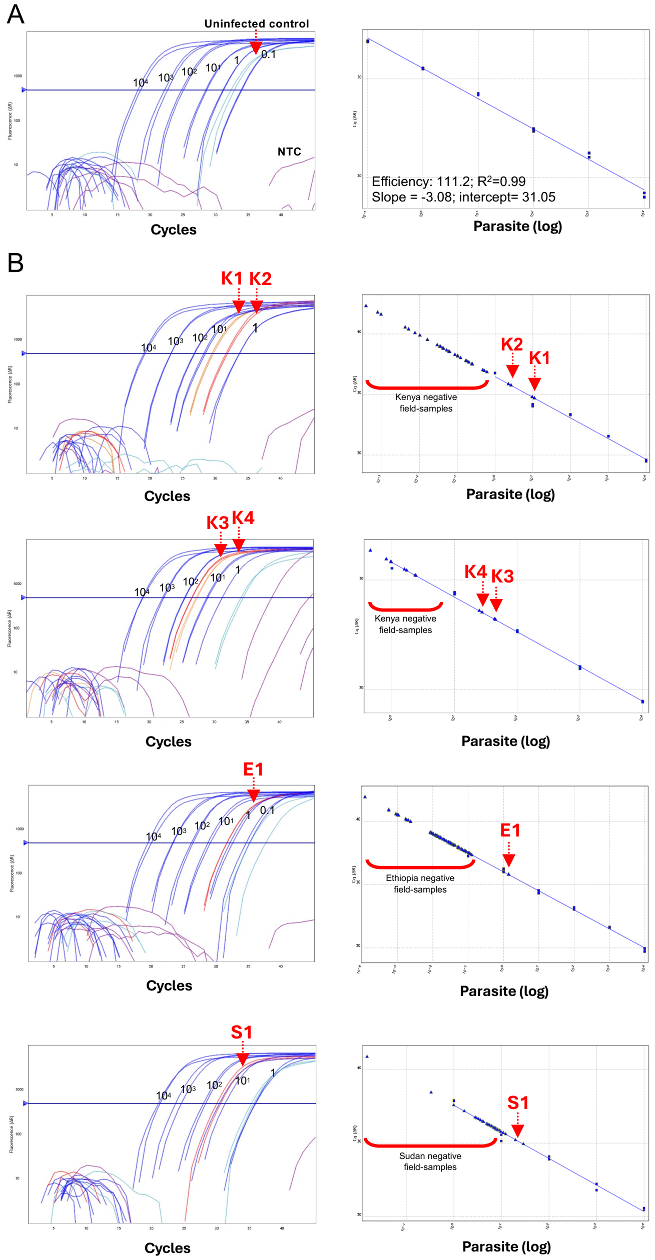


**Supplementary Figure 4. *Leishmania* qPCR on field-collected *Phlebotomus* sand flies**
(A) qPCR standard curve. (B) Representative qPCR amplification plots of positive *Leishmania*-infected field-collected samples from Kenya, Ethiopia, and Sudan. K, Kenya; E, Ethiopia; S, Sudan. Dark blue, standard curve from $10^4$ to 1 *Leishmania donovani* parasites spiked in one uninfected sand fly midgut (Kenya and Ethiopia) or whole body (Sudan); Aqua, uninfected negative sand fly midgut; purple, non-template control; Red or orange, field samples.

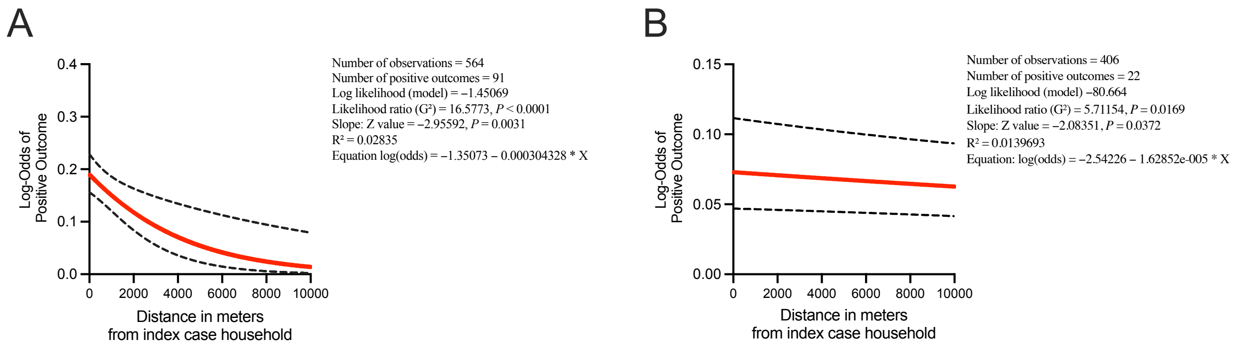


**Supplementary Figure 5. Effect of distance from index case household on rK39 seropositivity.** (A-B) Univariable binary logistic regression analyses was performed to assess the effect of distance (meters) from index case households to neighboring or control households, in correlation with rK39 positivity after normalization in Marsabit, Kenya (A), and Gedaref, Sudan (B). The ‘margins’ command of Stata was used to compute adjusted log-odds of seropositivity at selected distances from the index households. Analysis does not include index household members.

**Supplementary Table 1. Trapping sites and focality of *Leishmania*-infected *Phlebotomus* sand flies.**

| Country | Study site | Date of collection | Per trip | | | |
|---|---|---|---|---|---|---|
| | | | Average no. of traps per day | No. of traps with infected specimens | No. of infected specimens per trap | Trap ID |
| Marsabit Kenya | Laisamis | January, 2024 | 20 | 0 | 0 | - |
| | | April, 2024 | 20 | 4 | $1_i$, $1_i$, $1_i$, $3_i$ | 2, 10, 5, 4 |
| | | August, 2024 | 20 | 0 | 0 | - |
| | Log logo | January, 2024 | 20 | 0 | 0 | - |
| | | March, 2024 | 20 | 3 | $1_i$, $1_i$, $2_i$, $1_i$, $1_i$ | **3**, 8, **7**, **7**, **3** |
| | | July, 2024 | 20 | 10 | $3_p$+$1_i$, $1_p$, $1_i$, $1_i$, $1_i$, $1_i$, $1_i$, $1_i$, $1_i$, $1_i$, $1_p$, | 19, 1, 17, 13, **2**, 7, **2**, 21, 22, 11, 5 |
| | | August, 2024 | 20 | 0 | 0 | - |
| Gedaref state Sudan | Kunaynah | September, 2023 | 12 | 0 | 0 | - |
| | | October, 2023 | 12 | 0 | 0 | - |
| | | December, 2023 | 12 | 2 | $1_i$, $1_i$ | 5, 7 |
| | | January, 2024 | 12 | 2 | $1_p$, $1_i$ | 10, 8 |
| | | September, 2024 | 12 | 0 | 0 | |
| | Umslala-Houng and Dinder Park | November, 2023 | 12 | 0 | 0 | - |
| | | April, 2024 | 12 | 0 | 0 | - |
| | | June, 2024 | 12 | 5 | $1_i$, $2_i$, $1_i$, $1_i$, $1_i$ | 6, 7, 10, 8, 12 |
| Konso Aba-Roba Ethiopia | Galga | September, 2023 | 9.5 | 1 | $1_i$ | **14** |
| | | November, 2023 | 9.5 | 0 | 0 | - |
| | | February, 2024 | 12 | 0 | 0 | - |
| | | June, 2024 | 15 | 2 | $1_i$, $1_i$ | **14**, 15 |
| | Maira | September, 2023 | 7 | 0 | 0 | - |
| | | November, 2023 | 10.33 | 0 | 0 | - |
| | | February, 2024 | 11 | 0 | 0 | - |
| | | June, 2024 | 15 | 0 | 0 | - |
| | Lahalaha and Goinada | September, 2023 | 15 | 0 | 0 | - |
| | | November, 2023 | 10.33 | 0 | 0 | - |
| | | February, 2024 | 11 | 0 | 0 | - |

Specimens processed individually (i) or as a pool (p). D, day.Bold, trap IDs that appear more than once per study site.

**Supplementary Table 2. Primer sequences and concentrations used in the custom-designed multiplex panel for host blood meal identification by PCR and for generic *cytb* gene amplification for sequencing.**

| Host-blood meal multiplex-PCR target species | | | | | |
|---|---|---|---|---|---|
| **Species** | **5'-3' Sequence** | **Band Size (bp)** | **Stock μM** | **Reaction μM** | **μl per primer for a 30μL reaction** |
| Mongoose Fwd* | GGTGAAACCTCGGCTCTCTT | 937 | 10 | 0.183 | 0.55 |
| Equine (donkey/horse) Fwd* | ACTGCCTTCTCATCCGTCAC | 845 | 10 | 0.117 | 0.35 |
| Chicken Fwd | CAGCAGACACATCCCTAGCC | 723 | 10 | 0.042 | 0.125 |
| Chicken Rev | AGGCTGCTAGGGCTAGTACA | 723 | 10 | 0.042 | 0.125 |
| Camel Fwd* | GGCTACGTCCTTCCATGAGG | 638 | 10 | 0.033 | 0.1 |
| Ovis (goat/sheep) Fwd* | TGGCACAAACCTAGTCGAATGA | 558 | 10 | 0.067 | 0.2 |
| Dog Fwd* | CTCCCTTTCATCATCGCAGCTC | 473 | 10 | 0.05 | 0.15 |
| Hyrax Fwd* | CCAACGCCGACAAAATCCCAT | 388 | 10 | 0.133 | 0.4 |
| Cow Fwd* | AACTACACCCCAGCCAATCC | 263 | 10 | 0.05 | 0.15 |
| Human Fwd[42] | TTCGGCGCATGAGCTGGAGTCC | 228 | 10 | 0.067 | 0.2 |
| Human Rev[42] | TATGCGGGGAAACGCCATATCG | 228 | 10 | 0.067 | 0.2 |
| Muroidea Fwd[43] | TCTGGTTCTTACTTCAGGGC | 175 | 20 | 0.5 | 0.75 |
| Muroidea Rev[43] | TTCATGCCTTGACGGCTATG | 175 | 20 | 0.5 | 0.75 |
| Vertebrate UNRev[41]* | GGTTGTCCTCCAATTCATGTTA | - | 10 | 0.267 | 0.8 |
| Vertebrate AltRev* | GGCTGGCCTCCAATTCATGTGA | - | 10 | 0.033 | 0.1 |
| **Generic vertebrate *Cytb* primer sequence** | | | | | |
| CytB Fwd | CCATCAAACATCTCAGCATGATGAAA | 358 | 10 | 0.5 | 1.25 |
| CytB Rev | CCCCTCAGAATGATATTTGTCCTC | 358 | 10 | 0.5 | 1.25 |

*Forward primer for use with listed universal vertebrate reverse primer.

**Supplementary Table 3. Date of collection and ecotype of infected *Phlebotomus* sand flies from our study sites.**

<table>
<tr><th>Study site</th><th>Peridomestic /sylvatic</th><th>Ecotype</th><th>Date of collection</th><th>Physiological status, blood meal source</th></tr>
<tr><td rowspan="20">Log Logo, Kenya</td><td rowspan="16">Sylvatic</td><td>Vegetation</td><td>3/27/24</td><td>Unfed</td></tr>
<tr><td>Vegatation; vertisols</td><td>3/28/24</td><td>Unfed</td></tr>
<tr><td>Vegatation; vertisols</td><td>3/29/24</td><td>Unfed</td></tr>
<tr><td>Vegatation; vertisols</td><td>3/29/24</td><td>Unfed</td></tr>
<tr><td>Vegatation; vertisols</td><td>3/31/24</td><td>Unfed</td></tr>
<tr><td>Vegetation</td><td>3/31/24</td><td>Unfed</td></tr>
<tr><td>Vegetation</td><td>07/24/24</td><td>Blood fed, human</td></tr>
<tr><td>Vegatation; vertisols</td><td>07/24/24</td><td>Blood fed, rodent</td></tr>
<tr><td>Vegetation</td><td>07/24/24</td><td>Blood fed, human</td></tr>
<tr><td>Vegatation; vertisols</td><td>07/24/24</td><td>Blood fed, Gazelle</td></tr>
<tr><td>Vegatation; vertisols</td><td>07/24/24</td><td>Unfed pool</td></tr>
<tr><td>Vegatation; vertisols</td><td>07/24/24</td><td>Unfed pool</td></tr>
<tr><td>Vegatation; vertisols</td><td>07/24/24</td><td>Unfed pool</td></tr>
<tr><td>Animal enclosure</td><td>07/24/24</td><td>Unfed pool</td></tr>
<tr><td>Vegetation</td><td>07/24/24</td><td>Gravid</td></tr>
<tr><td>Temporary shelther</td><td>07/26/24</td><td>Blood fed, human</td></tr>
<tr><td rowspan="4">Peridomestic</td><td>Animal enclosure</td><td>07/26/24</td><td>Blood fed, human</td></tr>
<tr><td>Outside house</td><td>07/26/24</td><td>Blood fed, Gazelle</td></tr>
<tr><td>Animal enclosure</td><td>07/26/24</td><td>Blood fed, Gazelle</td></tr>
<tr><td>Indoors</td><td>07/29/24</td><td>Unfed pool</td></tr>
<tr><td rowspan="6">Laisamis, Kenya</td><td rowspan="6">Peridomestic</td><td>Outside house of VL patient #2, Patient relapsed in January, 2024; no treatment received; HIV* negative.</td><td>4/10/24</td><td>Gravid</td></tr>
<tr><td>Outside house of neighbor #5 of VL patient #2</td><td>4/10/24</td><td>Gravid</td></tr>
<tr><td>Inside house of neighbor #3 of VL patient #2</td><td>4/10/24</td><td>Unfed</td></tr>
<tr><td>Outside house of neighbor #2 of VL patient #2</td><td>4/11/24</td><td>Unfed</td></tr>
<tr><td>Outside house of neighbor #2 of VL patient #2</td><td>4/11/24</td><td>Gravid</td></tr>
<tr><td>Outside house of neighbor #2 of VL patient #2</td><td>4/11/24</td><td>Unfed</td></tr>
<tr><td rowspan="4">Kunaynah, Sudan</td><td rowspan="4">Peridomestic</td><td>Inside house of VL patient #3</td><td>12/15/23</td><td>Blood fed, human</td></tr>
<tr><td>Inside house of VL patient #3's neighbor</td><td>12/16/23</td><td>Blood fed, human</td></tr>
<tr><td>Inside house</td><td>1/7/24</td><td>Blood fed, unkown</td></tr>
<tr><td>Inside house</td><td>1/7/24</td><td>Unfed pool</td></tr>
<tr><td rowspan="4">Umslala-Houg & Dinder Park, Sudan</td><td rowspan="4">Peridomestic</td><td>Outside house</td><td>6/7/24</td><td>Blood fed, human</td></tr>
<tr><td>Animal enclosure</td><td>6/7/24</td><td>Blood fed, human</td></tr>
<tr><td>Animal enclosure</td><td>6/7/24</td><td>Blood fed, human</td></tr>
<tr><td>Outside house</td><td>6/7/24</td><td>Blood fed, human</td></tr>
</table>

| | | | | |
|---|---|---|---|---|
| | Sylvatic | Termite hill | 06/11/24 | Blood fed, human |
| | | Vegetation | 06/13/24 | Blood fed, human |
| Galga, Ethiopia | Peridomestic | Termite hill | 9/27/23 | Unfed |
| | | Termite hill | 06/23/24 | Gravid |
| | | Termite hill | 06/23/24 | Gravid |

* Human immunodeficiency virus.

**Supplementary Table 4. Confirmation of blood meal host identification of *Leishmania* infected samples by cytochrome b (*cyt b*) gene sequencing.** PCR amplification of vertebrate *cytb* gene using universal primer sets was carried out and the product was sequenced. BLAST results confirmed the host with exact or nearly exact matches by the percent identity. A bit E-score >50, coverage >35%, identity >70% were considered as valid.

| Study site | Species by *Cyt b* sequencing | Query Cover primer | E-value | % Identity | Species |
|---|---|---|---|---|---|
| Log Logo, Kenya | Gazelle | 79% | 2.00E-118 | 92.46% | Gazelle |
| Log Logo, Kenya | Human | 97% | 9.00E-158 | 99.38% | Human |
| Log Logo, Kenya | Human | 95% | 1.00E-150 | 98.44% | Human |
| Log Logo, Kenya | Human | 97% | 7.00E-153 | 98.46% | Human/Dog |
| Log Logo, Kenya | Human | 96% | 1.00E-156 | 99.07% | Human |
| Log Logo, Kenya | Gazelle | 83% | 3.00E-134 | 93.94% | Gazelle |
| Log Logo, Kenya | Gazelle | 93% | 2.00E-110 | 89.81% | Gazelle |
| Log Logo, Kenya | - | - | - | - | Rodent |
| Dinder Park, Sudan | Human | 85% | 1.00E-151 | 97.55% | Human |
| Dinder Park, Sudan | Human | 96% | 4.00E-156 | 98.77% | Human |
| Umslala-Houg, Sudan | Human | 97% | 2.00E-154 | 98.46% | Human |
| Umslala-Houg, Sudan | Human | 93% | 6.00E-129 | 93.91% | Human |
| Umslala-Houg, Sudan | Human | 97% | 3.00E-158 | 99.38% | Human |
| Umslala-Houg, Sudan | Human | 96% | 1.00E-157 | 99.38% | Human |
| Kunaynah, Sudan | Human | 68% | 4.00E-149 | 98.69% | Human |
| Kunaynah, Sudan | Human | 67% | 2.00E-142 | 97.69% | Human |
| Kunaynah, Sudan | Uknown | - | - | - | Unkown |

**Supplementary Table 5. Dates of diagnosis and treatment and serum collection from VL patients in our study sites in Marsabit county, Kenya, and Gedaref state, Sudan.**

| Study site | Patient ID | Date of clinical diagnosis (rK39 RTD and /or microscopy) and treatment | This study Date of DBS collection | This study DBS rK39 ELISA |
|---|---|---|---|---|
| Laisamis, Marsabit Kenya | 1 | December, 2023 | January, 2024 | - |
| | 2 | First diagnosis and treatment in 2021. Patient relapsed in January, 2024; no treatment received; HIV* negative. | April, 2024 | - |
| | 3 | February, 2024 | April, 2024 | - |
| Karare, Marsabit Kenya | 4 | July, 2024 | August, 2024 | + |
| | 5 | June, 2024 | August, 2024 | + |
| | 6 | June, 2024 | August, 2024 | - |
| | 7 | June, 2024 | August, 2024 | - |
| Laisamis, Marsabit Kenya | 8 | September, 2024 | January, 2025 | - |
| | 9 | September, 2024 | January, 2025 | - |
| | 10 | September, 2024 | January, 2025 | - |
| | 11 | October, 2024 | January, 2025 | - |
| | 12 | October, 2024 | January, 2025 | + |
| | 13 | November, 2024 | January, 2025 | - |
| | 14 | November, 2024 | January, 2025 | - |
| | 15 | November, 2024 | February, 2025 | + |
| | 16 | December, 2024 | February, 2025 | + |
| | 17 | December, 2024 | February, 2025 | + |
| | 18 | December, 2024 | February, 2025 | + |
| | 19 | December, 2024 | February, 2025 | - |
| | 20 | December, 2024 | February, 2025 | - |
| | 21 | January, 2025 | February, 2025 | - |

| | | | | |
|---|---|---|---|---|
| Laisamis, Marsabit Kenya | 22 | January, 2025 | February, 2025 | + |
| | 23 | January, 2025 | February, 2025 | + |
| | 24 | January, 2025 | February, 2025 | + |
| | 25 | January, 2025 | February, 2025 | + |
| | 26 | February, 2025 | February, 2025 | - |
| | 27 | February, 2025 | February, 2025 | - |
| | 28 | January, 2025 | May, 2025 | + |
| | 29 | January, 2025 | May, 2025 | + |
| | 30 | February, 2025 | May, 2025 | - |
| | 31 | February, 2025 | May, 2025 | - |
| | 32 | February, 2025 | May, 2025 | + |
| | 33 | March, 2025 | May, 2025 | + |
| | 34 | March, 2025 | May, 2025 | - |
| | 35 | March, 2025 | May, 2025 | + |
| | 36 | March, 2025 | May, 2025 | - |
| | 37 | March, 2025 | May, 2025 | - |
| | 38 | March, 2025 | May, 2025 | + |
| | 39 | March, 2025 | May, 2025 | + |
| | 40 | March, 2025 | May, 2025 | - |
| | 41 | April, 2025 | May, 2025 | - |
| | 42 | April, 2025 | May, 2025 | + |
| | 43 | April, 2025 | May, 2025 | - |
| | 44 | April, 2025 | May, 2025 | + |
| | 45 | April, 2025 | May, 2025 | + |
| | 46 | April, 2025 | May, 2025 | + |
| | 47 | April, 2025 | May, 2025 | - |
| | 48 | May, 2025 | May, 2025 | - |
| | 1 | August, 2023 | October, 2023 | - |

| | | | | |
|---|---|---|---|---|
| Kunaynah, Gedaref state Sudan | 2 | September, 2023 | December, 2023 | - |
| | 3 | November, 2023 | December, 2023 | - |
| | 4 | December, 2023 | January, 2024 | - |
| | 5 | August, 2024 | September, 2024 | - |

* Human immunodeficiency virus.
